# Pruning and thresholding approach for methylation risk scores in multi-ancestry populations

**DOI:** 10.1101/2022.06.09.22276204

**Authors:** Junyu Chen, Evan Gatev, Todd Everson, Karen N. Conneely, Nastassja Koen, Michael P. Epstein, Michael S. Kobor, Heather J. Zar, Dan J. Stein, Anke Huels

## Abstract

Recent efforts have focused on developing methylation risk scores (MRS), a weighted sum of the individual’s DNAm values of pre-selected CpG sites. Most of the current MRS approaches that utilize Epigenome-wide association studies (EWAS) summary statistics only include genome-wide significant CpG sites and do not consider co-methylation. New methods that relax the p-value threshold to include more CpG sites and account for the inter-correlation of DNAm might improve the predictive performance of MRS. We paired informed co-methylation pruning with P-value thresholding to generate pruning and thresholding (P+T) MRS and evaluated its performance among multi-ancestry populations. Through simulation studies and real data analyses, we demonstrated that pruning provides an improvement over simple thresholding methods for prediction of phenotypes. We demonstrated that European-derived summary statistics can be used to develop P+T MRS among other population such as African population. However, the prediction accuracy of P+T MRS may differ across multi-ancestry population due to environmental/cultural/social differences.

## Introduction

DNA methylation (DNAm), one of the most studied epigenetic mechanisms, regulates the mode of expression of DNA segments independent of alterations of their sequence by adding a methyl group at cytosine residues, hence contributing to variation in cellular phenotypes^1^. With current advances in and reduction of cost of array-based profiling technologies, increasing numbers of large-scale epigenome-wide association studies (EWAS) have been conducted to study DNAm in association with complex human diseases as well as environmental and social factors^2,3^. EWAS has thus far been successful in identifying dozens of cytosine guanine dinucleotides (CpGs) associated with various diseases and exposures, which could potentially be used for disease diagnosis and prediction, development of drug targets, and monitoring of drug response^3-8^. However, differential DNAm in individual CpGs often shows a weak prediction capacity and can only explain a small fraction of phenotype variance. Polyepigenetic approaches that aggregate information on differential DNAm from multiple CpGs might produce a more accurate biomarker for clinical usage^9,10^.

A well-known polygenic approach for genotype data is polygenic risk scores (PRS), which are weighted sum of risk alleles of a pre-selected number of genetic variants^11^. Recently, many efforts have focused on transferring PRS approaches to DNA methylation data to construct methylation risk scores (MRS), which are defined as weighted sums of the individuals’ DNAm values of a pre-selected number of CpGs^10^. However, there are many methodological challenges in constructing DNA methylation risk scores^10,12,13^. One of the problems is that DNAm is influenced by ancestry, which captures genetic ancestry (differences in the genome related to ancestry) as well as social determinants of health such as racism and discrimination, socioeconomic status, and environmental effects^14^. Thus, ideally, when external weights are used for the calculation of MRS, these weights should be assessed in a population with the same ancestry as the study samples. However, current epigenetic literature remains limited by the lack of diversity, with most focusing on European populations^15^, therefore making it difficult to identify appropriate weights for MRS for other populations. While it is well known that PRSs are not applicable across different ancestries^16,17^, little is known about the performance of MRS across multi-ancestry populations.

Currently, there are two popular approaches to construct MRS. The first one is to use penalized regression models such as Elastic Net and LASSO regularization^10,18,19^, which usually requires individual-level DNAm data. When only summary-level statistics are available, individual CpGs that reached genome-wide significance in an external epigenome-wide association studies (EWAS) are selected and the beta-coefficients estimated from EWAS are used as weights to generate MRS^10^. However, research in PRS has shown that the optimal p-value threshold strongly depends on the data^20^, and including a larger proportion of variants could potentially capture more of the phenotype variation^21^. Moreover, most MRS from the second approach do not consider DNA co-methylation, defined as proximal CpGs with correlated DNAm across individuals^22^, which could potentially bias the generation of MRS. Shah et al 2015 proposed to remove redundant CpGs by keeping the most significant CpGs in co-methylation^23,24^, however, a window to define DNA co-methylation needs to be pre-defined and the effect of accounting for DNA co-methylation was not evaluated.

One of the most widely used PRS approaches to deal with single nucleotide polymorphisms (SNPs) in high linkage disequilibrium (LD) and to identify p-value thresholds with the best prediction accuracy is the pruning and threshold (P+T) method^25^. In the P+T approach, the correlation square (R^2^) for SNPs within a close genetic distance is calculated and less significant SNPs that are correlated with an R^2^ greater than a particular value (LD pruning)^26^ are removed. Next, several p-value thresholds are tested to maximize the prediction accuracy of the derived PRS (p-value thresholding)^26,27^. Theoretically, the P + T approach could be applied to generate MRS, however, there is no standard procedure on how to conduct pruning for DNAm data and the performance of such MRS across multi-ancestry populations remains unknown.

Here, we propose to use the Co-Methylation with genomic CpG Background (CoMeBack), a tool that uses a sliding window to estimate DNA co-methylation, to account for correlations of DNAm at proximal CpG sites^22^, and pair it with p-value thresholding to construct P+T CoMeBack MRS. CoMeBack uses unmeasured intermittent CpGs from the human reference genome to link array probes in hope of reducing false positives while improving the identification of biologically relevant co-methylation^22^. We conducted simulation studies based on data from an adult population consisting of three groups of different ancestries (Indian, White and Black, n = 1,199) to evaluate the prediction performance of P+T CoMeBack MRS and how it changes across multi-ancestry population. Next, we applied the P+T CoMeBack approach to DNAm data from the Drakenstein Child Health Study (n=270)^28^, a multi-ancestry birth cohort from South Africa, to evaluate the performance of MRS for maternal smoking status. Our simulation study and real data application demonstrated that the P+T approach improves the predictive accuracy of MRS over methods that do not account for co-methylation and has similar performance as LASSO regression, which requires access to the raw DNAm data. We also showed that MRS built upon the data from a population of one genetic ancestry could achieve high prediction performance among populations of other genetic ancestries, but the performance might differ in the presence of environmental/cultural/social differences associated with ancestry.

## Materials and methods

### P+T CoMeBack approach for MRS

P+T CoMeBack method refers to the calculation of MRS using informed co-methylation pruning (P) with CoMeBack and P-value thresholding (T). First, summary statistics from an EWAS (typically include the participant ID, effect size, standard error and P-value of each CpG site) need to be estimated in an independent dataset (training dataset) to avoid overfitting, and then applied to generate MRS in a testing dataset (the samples used to evaluate the performance of MRS).

In our P+T CoMeBack method, co-methylation pruning is completed by applying CoMeBack to DNAm data of the testing dataset or a reference panel^22^. Specifically, CoMeBack chains two adjacent array probes if the following requirements are met: 1. two probes are less than 2kb apart; 2. the reference human genome annotation shows a set of unmeasured genomic CpGs between them; 3. the density of unmeasured genomic CpGs between them is at least one CpG every 400bp. Chaining of adjacent array probes continues until an array probe does not meet the requirements, which will form a unit where multiple CpGs are chained together. Correlations between DNAm levels will then be calculated for all array probes inside each unit. If all pairs of adjacent probes in a unit have a correlation square (R^2^) greater than 0.3, such unit will be declared as a co-methylated region (CMR). Pruning is conducted by only keeping one CpG site per CMR in the dataset, the one with the lowest (most significant) P-value in the EWAS summary statistics.

P + T CoMeBack will be compared to the standard pruning approach, in which less significant SNPs that are correlated with an R^2^ > 0.3 and located within 2000bp of each other are being removed.

Next, P-value thresholding step (T) is performed for the pruned set of CpG sites. Specifically, the P-value thresholding step (T) is performed by applying different P-value thresholds (e.g., P-value thresholds ∈ [0.05, 0.005, 5 × 10^−4^, 5 × 10^−5^, …]) and only including those CpG sites in the final MRS calculation that reached a P-value below those thresholds in the EWAS summary statistics.

Finally, for each P-value threshold, MRS are calculated as a weighted sum of DNAm *β* values (*β* value = methylated allele intensity / (unmethylated allele intensity + methylated allele intensity + 100), ranging from 0 representing unmethylation to 1 for complete methylation) of the selected CpGs, where the weights are the corresponding effect sizes for each CpG from the EWAS summary statistics. The squared correlation (R^2^) between the phenotype of interest and MRS obtained using each P-value threshold is calculated to represent the prediction accuracy. The P-value threshold that produces MRS with the highest prediction accuracy in the testing data set is selected as the optimal P-value and the corresponding MRS is used for downstream analysis. The pipeline for generating P+T MRS is written in an R script, which is available at GitHub (https://github.com/jche453/Pruning-Thresholding-MRS.git).

In our simulation studies and real data application, we compare the P+T CoMeBack MRS approach to the standard P+T and T approach, which refers to an approach in which the MRS is calculated by only thresholding, not accounting for correlations between included CpGs (no pruning).

### Simulation studies

To validate the performance of the proposed P+T MRS approach, we conducted simulation studies based on whole blood Illumina Infinium Human Methylation 450K BeadChip data from an ethnically heterogeneous discovery cohort composed of several publicly available datasets (GSE55763, GSE84727, GSE80417, GSE111629 and GSE72680)^22^. Intra-dataset normalization and batch effects correction were performed using ComeBat function in R-package sva^29^, followed by merging of datasets and correction for inter-dataset batch effects using the same function. After the removing XY chromosome binding, non-CpG, cross-hybridizing probes and probes that are in close distance with common SNPs, there were 386,362 CpGs left for MRS analysis. We randomly selected 1,199 adults (898 Indians, 136 Blacks and 165 Whites) to conduct the simulation studies.

CoMeBack was applied to the DNA methylation *β* values of the 386,362 CpGs to obtain CMR. In each simulation, 10 of the 386,362 CpGs were randomly selected to be causal, k% (k = 30, 50, 70 or 100) of which are in a CMR with other CpGs. At most, one CpG would be causal in each CMR.

The causal CpGs were randomly assigned a “true” effect size from a uniform distribution as *w*_*i*_ ∼ *U*(−0.5, 0.5). We then simulated a phenotype for the *j-*th subject as follow:

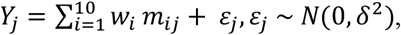

where *m*_*ij*_ is the DNAm *β* value of causal CpG site *i* of the *j-*th subject, and *ε*_*j*_ is an error term that follows a normal distribution. Different *δ*^2^ were set to ensure that the targeted variance of phenotype explained by DNAm alone equals 10%, 30% 50% or 80%.

We also simulated a second phenotype Y*_j_ fjor the *j*-th subject, which was directly affected by ancestry using European ancestry as reference:

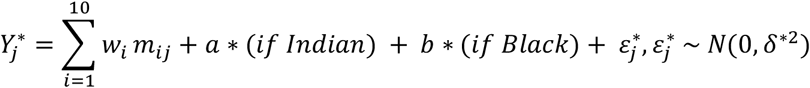

Effect of ancestry in our simulations is simulated as the effect of genetic ancestry assuming there were no complex social determinants involved in the causal pathway. Different *δ*\*^2^ were used so that the variance of phenotype that was explained by DNAm and ancestry together equals 20%, 50% or 80. For our simulations, effect *a* was set to 0.1 and *b* to 0.2. In each simulation, both simulated phenotypes *Y*_*j*_ and 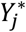 share the same epigenetic liability 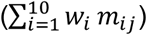.

In each simulation, for fair comparison, 762 Indians were randomly chosen as the training dataset so that there were at least 136 people left for each race group in the testing dataset. Associations between CpGs and each of the two simulated phenotypes were assessed by robust linear regression model using limma R package^30^ in the training dataset. We calculated top 10 principal components (PCs) from DNAm of 386,362 CpGs^31^ and used EpiDISH to estimate cell type proportions of each CpGs^32^. We observed that in our simulation dataset, top 10 PCs are highly correlated with cell type proportions (Supplement figure 1A), and using either summary statistics adjustment for top 10 PCs or summary statistics adjusted for cell type proportions would lead to almost identical prediction performance of MRS (Supplement figure 1B). Thus, to account for population stratification and cell type difference, we adjusted for the top 10 PCs in our main analyses. The summary statistics (effect size and P-values) obtained from association tests in the training data were saved and later used to construct MRS in the testing dataset. We repeated 1000 simulations per scenario to evaluate the prediction accuracy (R^2^), power and type 1 error rate of the P+T MRS. Linear regression analysis was used to access the association between MRS and simulated phenotypes, and power is defined as the proportion of simulations where MRS were significantly associated with the simulated phenotype with at α level of 0.05. To estimate type 1 error rate, we first obtained a null association between MRS values and simulated phenotype values by permutation of MRS values. Linear regression analysis was used to access the association between permutated MRS and simulated phenotypes, and type 1 error rate is defined as the proportion of simulations where permutated MRS were significantly associated with the simulated phenotype.

We evaluated the performance of the MRS not only in scenarios of A) same ancestry in training and test data, but also B) across different ancestry groups (training data: Indian, test data: European or African) and C) in multi-ancestry populations (training data: Indian, test data: Indian, European and African). For scenario C), we evaluated two analysis strategies: 1. Joint-analysis: perform MRS analyses in the whole testing dataset where subjects from all racial groups were merged; 2. Standardization: scale MRS to have a standard normal distribution within each racial group before merging all subjects for analyses.

### Application study of smoking MRS

To evaluate the performance of the P+T CoMeBack approach in a real data setting, we applied the P+T CoMeBack approach to calculate a MRS for maternal smoking status during pregnancy using cord blood DNAm data from newborns in the South African Drakenstein Child Health Study (DCHS), a multi-ancestry longitudinal study investigating determinants of early child development^33^. There were 145 Black African infants and 115 Mixed ancestry infants in the DCHS. A detailed description of the enrollment process, inclusion criteria, variables measurement and ethical approval of the study have been previously published^33,34^.

Cotinine levels were measured in urine provided by mothers within four weeks of enrollment and classified as <499 ng/ml (non-smoker), or ≥500 ng/ml (active smoker)^28^. Cord blood was collected at time of delivery and used to measure DNA methylation by either MethylationEPIC BeadChips (EPIC, n=145) or the Illumina Infinium HumanMethylation450 BeadChips (450K, n=103)^33,34^, followed by quality control and normalization to calculate β values (details have been published elsewhere) ^35^.

Summary statistics for the calculation of MRS were obtained from a study that meta-analyzed the associations between newborn blood DNA methylation and sustained maternal smoking during pregnancy among 5,648 mother-child pairs as part of the Pregnancy and Childhood Epigenetics (PACE) Consortium (Table 1)^36^. The participants of all cohorts used in the meta-analysis except one were of European ancestry.

**Table 1.**
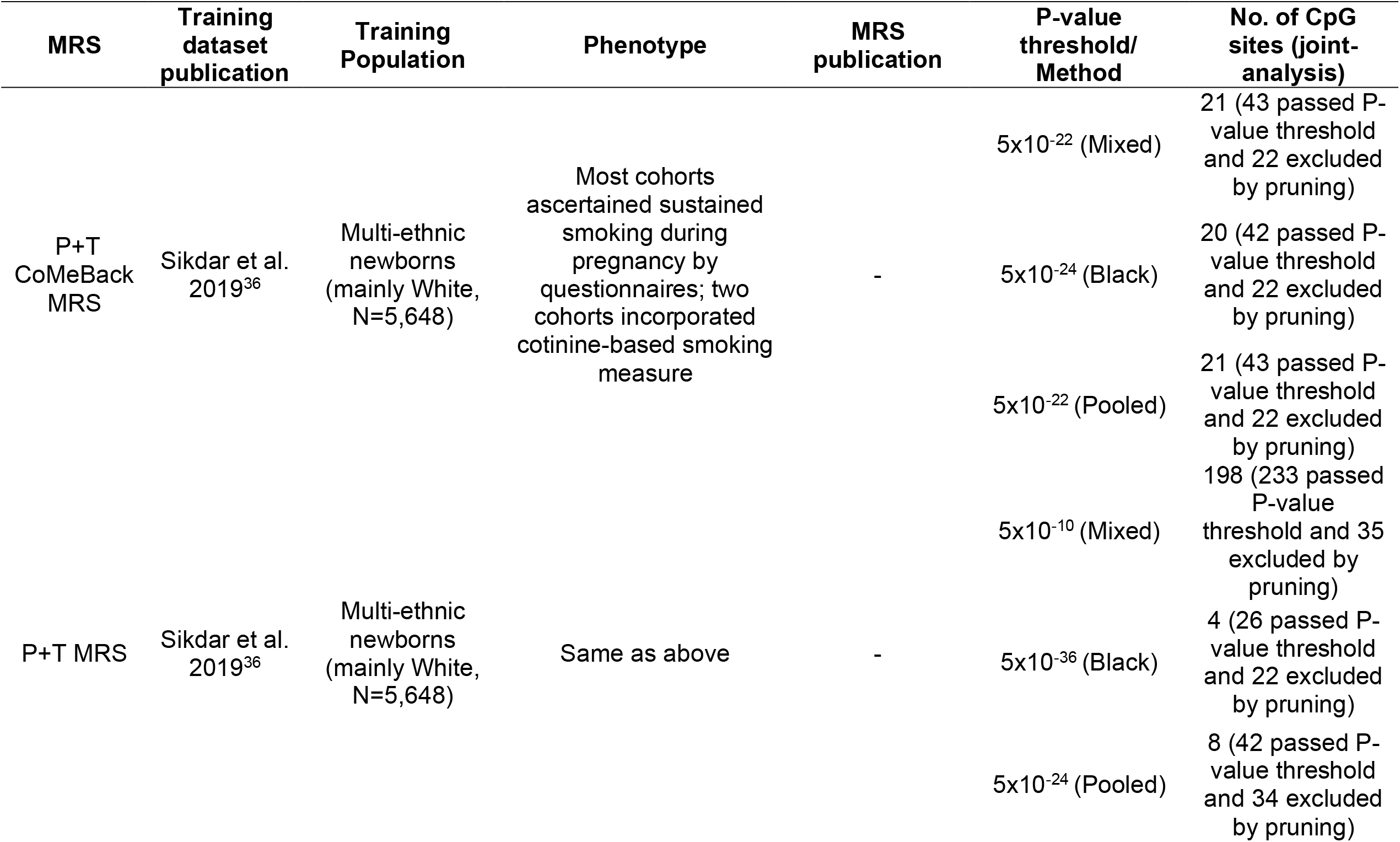
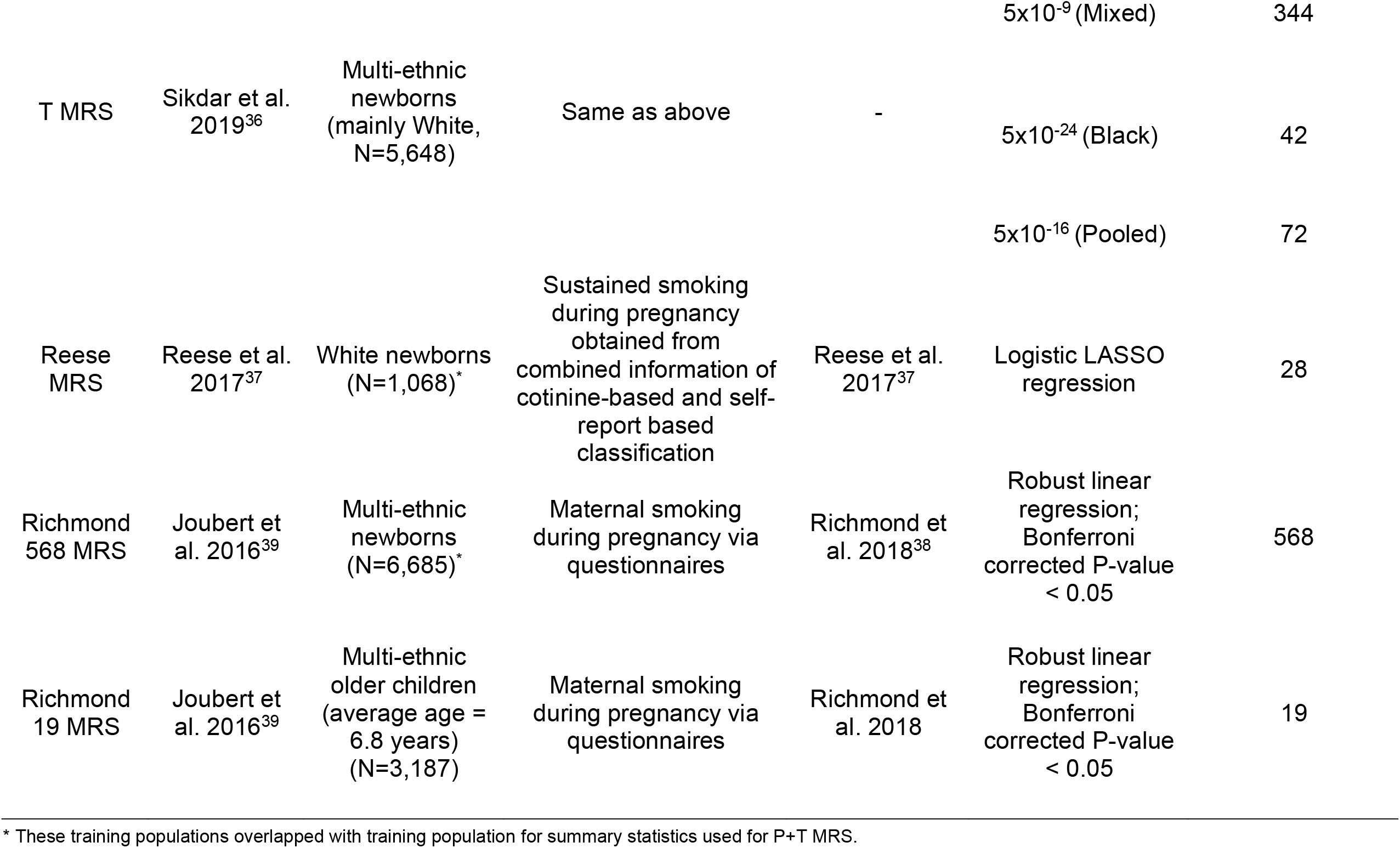
Overview of included EWAS, their phenotypes, training sample and methods.

In addition, we compared P+T CoMeBack MRS to three previously published MRS for maternal smoking during pregnancy (Reese MRS, Richmond 19 MRS, Richmond 568 MRS; Table 1). Reese MRS model was trained among 1,068 newborns of European ancestry in the Norwegian Mother and Child Cohort Study, while Richmond 568 MRS and Richmond 19 MRS was trained in multi-ancestry newborns (N=6,685) and children around 6.8 years old (N=3,187) in PACE Consortium respectively. The training population for Reese MRS and Richmond 19 MRS overlapped with the training population for summary statistics used in P+T CoMeBack MRS in our study. Reese et al. used a LASSO regression to select CpGs for Reese MRS, which is a weighted sum of DNAm β values of 28 CpGs with weights estimated from the LASSO regression^37^. Richmond 19 MRS is a weighted sum of DNAm β values of 19 CpGs that were significantly associated with prenatal smoking in an EWAS conducted in peripheral blood from children of averaged 6.8 years age (Richmond 19 MRS)^38,39^. In the same study, Richmond 568 MRS was proposed based on 568 CpGs that were significantly associated with prenatal smoking in cord blood^38,39^. We obtained the weights of reported CpGs from the mentioned studies and applied them to DNAm data in DCHS to generate Reese MRS, Richmond 19 MRS and Richmond 568 MRS.

Linear regressions were used to assess the associations between maternal smoking status and each MRS, controlling for ancestry (in pooled samples), cell type proportions and top 5 PCs calculated from genotypes. In order to obtain comparable beta-coefficients and standard errors across different MRS, each MRS was divided by their interquartile range (IQR) before linear regression analysis.

## Results

### Simulation results

We compared the prediction performance of P+T CoMeBack MRS to the T method among 136 Indians in the test data across different simulation scenarios (Figure 1). Figure 1A shows that P+T CoMeBack MRS that account for co-methylation between CpGs have stable prediction performance when proportion of causal CpGs located in a CMR (k%) varies. P+T without CoMeBack had similar prediction performance while the T method had a slightly lower prediction performance. While the P+T CoMeBack MRS showed subtle improvement over T method when 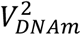 is 80% (Figure 1A), the difference between P+T CoMeBack MRS and the T method decreases as the 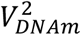 decreases. This is likely because as variance explained by DNAm decreases, there is less power for association testing, and it becomes increasingly difficult to distinguish real signals from statistical noise while generating the summary statistics.

**Figure 1.**
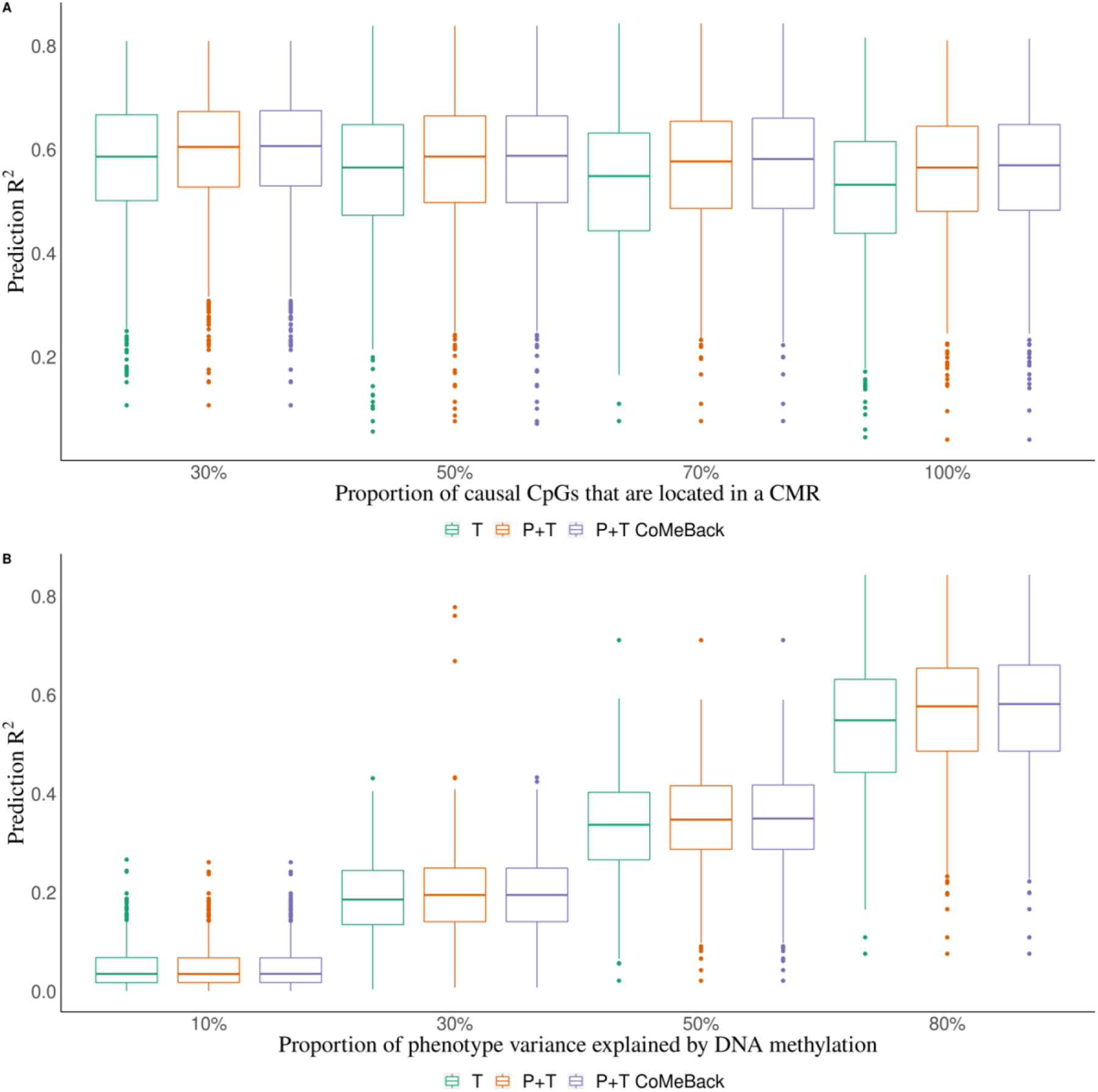
Simulation study. Prediction R^2^ of P+T CoMeBack, P+T and T method in dependence of (A) the proportion of causal CpG sites in CMRs and (B) proportion of phenotype variance explained by DNA methylation, among Indian participants. For each simulation, the discovery cohort was repeatedly and randomly split into a training set comprising 762 Indians and a testing set comprising 136 people of the same ancestry. Phenotypes were simulated without an influence of ancestry. Results are shown for (**A**) different proportions of causal CpGs located in CMR (30%, 50%, 70%, 100%) and (**B**) different proportions of phenotype variance explained by DNA methylation (10%, 30%, 50%, 80%). Each box represents the distribution of prediction accuracy across 1000 simulations, where the central mark is the median and the edges of the box are the 25^th^ and 75^th^ percentiles.

Next, we assessed the performance of P+T CoMeBack, P+T and T method across different ancestries and among multi-ancestry populations. All three methods achieved a high power (> 95%) and a low type 1 error rate (∼ 5%) within each ancestry in most scenarios for both phenotypes except when the phenotype variance explained by DNA methylation is 10% or 30% (Supplement table 1-4).

Whether the simulated phenotypes were independent of ancestry or not, MRS among Whites and Blacks achieved a prediction R^2^ as high as among Indians, which should have the best prediction of the simulated phenotypes since weights were obtained from Indian training samples (Figure 2). Findings were similar when the phenotype variance explained by DNA methylation was reduced from 80% to 10%, 30% or 50% (Supplement Figure 2). When the phenotypes are not associated with ancestry (Figure 2A), the three MRS analyses strategies (stratification, joint analysis and standardization) lead to nearly identical results. However, when the phenotypes are ancestry-dependent, both joint analysis and standardization of MRS showed very poor prediction of the phenotypes (Figure 2B).

**Figure 2.**
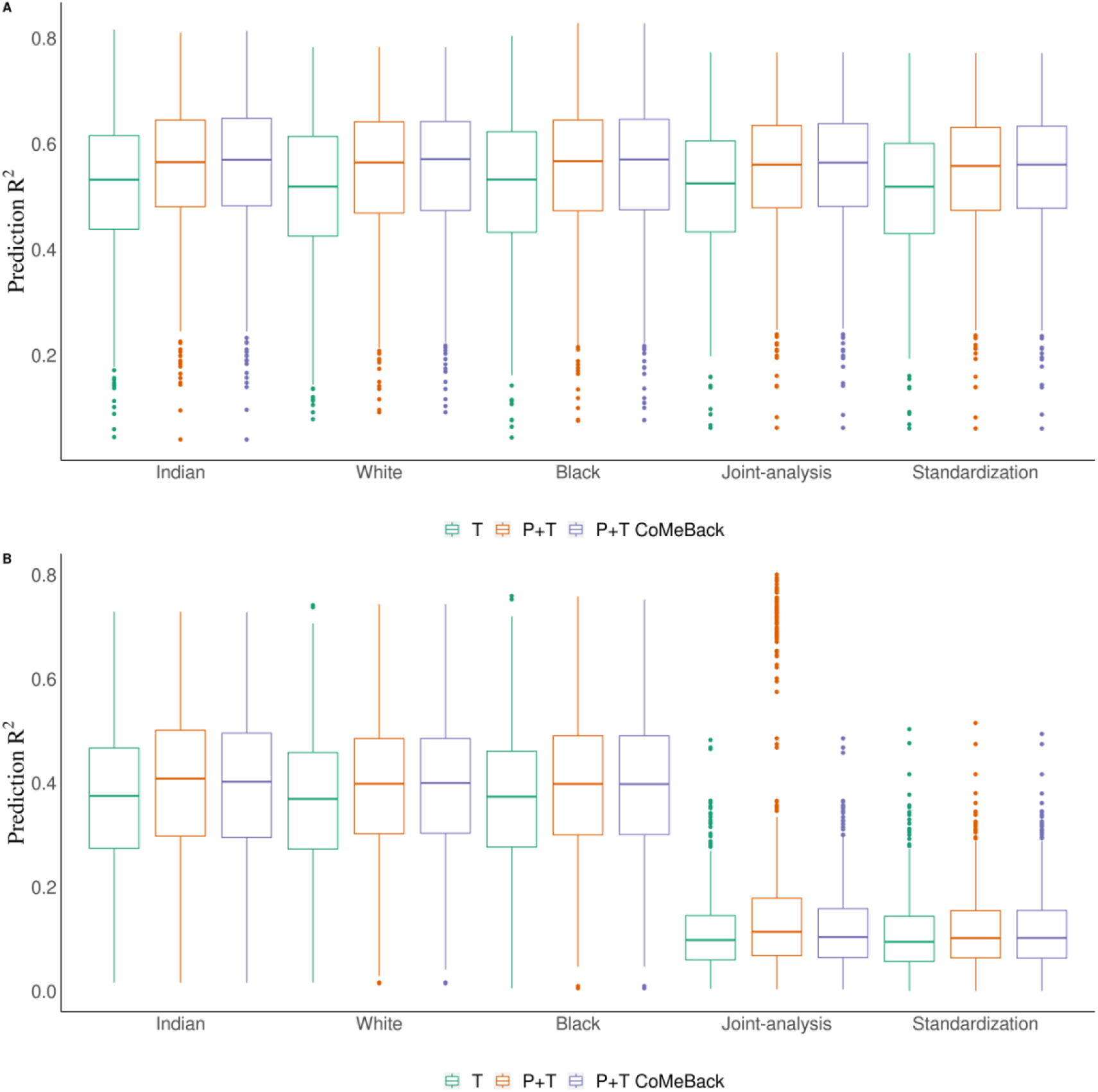
Simulation study. Prediction R^2^ of P+T CoMeBack and T approach across different racial groups and among multi-ancestry populations. For each simulation, the discovery cohort was repeatedly and randomly split into a training set comprising 762 Indians and a testing set comprising 136 people of each ancestry group. The proportion of causal CpGs located in CMR is 70% and the proportion of phenotype variance explained by DNA methylation (and ancestry) is 80%. Results are shown for the prediction of simulated phenotypes (**2A**) without an influence of ancestry and (**2B**) influenced by ancestry. Joint-analysis refers to MRS analyses of all participants pooled from all ancestry groups and standardization refers to standardizing MRS within each ancestry group and then merging all participants before analyses. Each box represents the distribution of prediction accuracy across 1000 simulations, where the central mark is the median and the edges of the box are the 25^th^ and 75^th^ percentiles.

### MRS of maternal smoking status

Figure 3 shows the prediction performance of MRS for maternal smoking status among DCHS newborns. As the p-value threshold decreases, the prediction accuracy of the resulting MRS increases before reaching a plateau, demonstrating the importance of P-value thresholding in MRS to control for noise. Among mixed ancestry newborns, P+T CoMeBack MRS of smoking status excluded 22 CpGs in pruning and achieved a prediction R^2^ of 29.5% using P-value threshold of 5×10^−22^, while the standard P+T without CoMeBack had a lower prediction R^2^ of 26.2% using P-value threshold of 5×10^−10^ and the best T method MRS had the lowest prediction accuracy (24.5%) using P-value threshold 5×10^−9^, confirming the benefits of pruning in MRS calculation (Figure 3A). All three MRS had lower prediction performance for maternal smoking among Black African infants (10.9%, and 8.0% respectively) (Figure 3B), which is likely due to the low prevalence of smokers among mothers of Black African infants in DCHS (13%) compared to mothers of mixed ancestry infants (49%) (Supplement Figure 3). Additionally, the distributions of all MRS in Black African infants and mixed ancestry infants were similar within each category of maternal smoking status (Supplement Figure 4), confirming that the difference of prediction R^2^ between Mixed and Black infants is less likely due to ancestry-related factors other than prevalence of maternal smoking. Joint-analysis of P+T CoMeBack MRS showed a prediction accuracy of 20.4%, which is between the prediction accuracy of P+T CoMeBack MRS among Black African infants and mixed ancestry infants (Figure 3C). Standardization approach did not improve the performance of MRS (Figure 3D).

**Figure 3.**
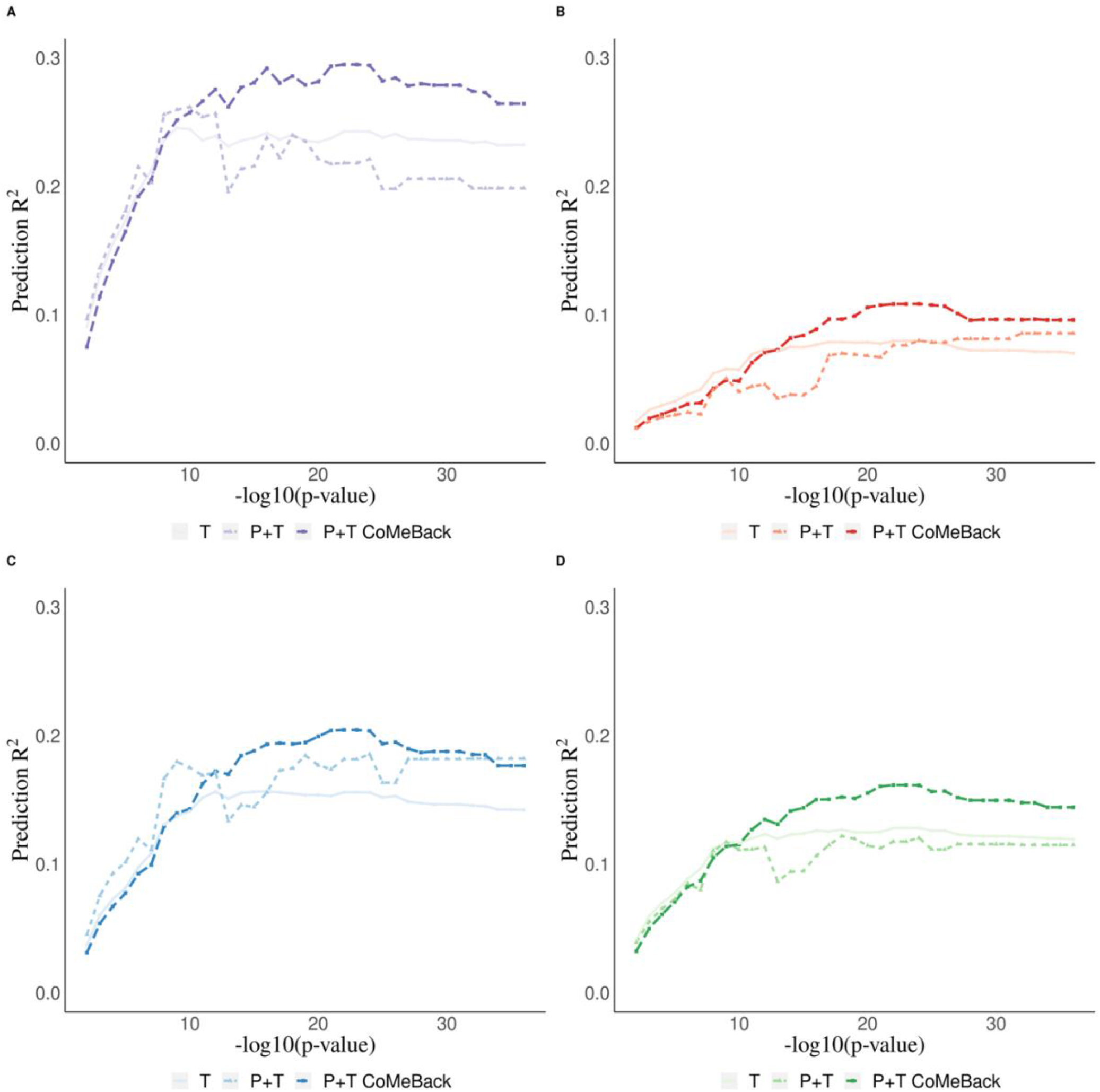
Real data application. MRS for the prediction of maternal smoking during pregnancy using cord blood DNA methylation data from newborns in the South African Drakenstein Child Health Study (DCHS). Prediction R^2^ of maternal smoking status is shown stratified for **A**. Mixed infants. **B**. Black infants. **C**. joint-analysis (all subjects pooled from all ancestries) **D**. Standardization (standardizing MRS within each ancestry and merging all subjects before analyses

We next compared the prediction accuracy and distribution of P+T CoMeBack MRS to other established MRS for maternal smoking during pregnancy and newborn DNAm (Figure 4). Overall, P+T CoMeBack and Reese MRS had stable and similar classification performance in all analyses compared to other MRS. P+T CoMeBack MRS and Reese MRS showed a similar prediction R^2^ among both Black and Mixed ancestry infants, which are better than other smoking MRS (Figure 4A). P+T CoMeBack MRS had the largest AUC (0.820) in the ROC curve among mixed infants (Figure 4B) but a smaller AUC than Reese MRS in Black infants and joint-analysis (Figure 4C-D). Further, all 6 MRS showed significant association with smoking status in Mixed infants, Black infants and joint-analysis (Table 2), showing the promise of using MRS to capture the overall DNAm signals in association testing.

**Figure 4.**
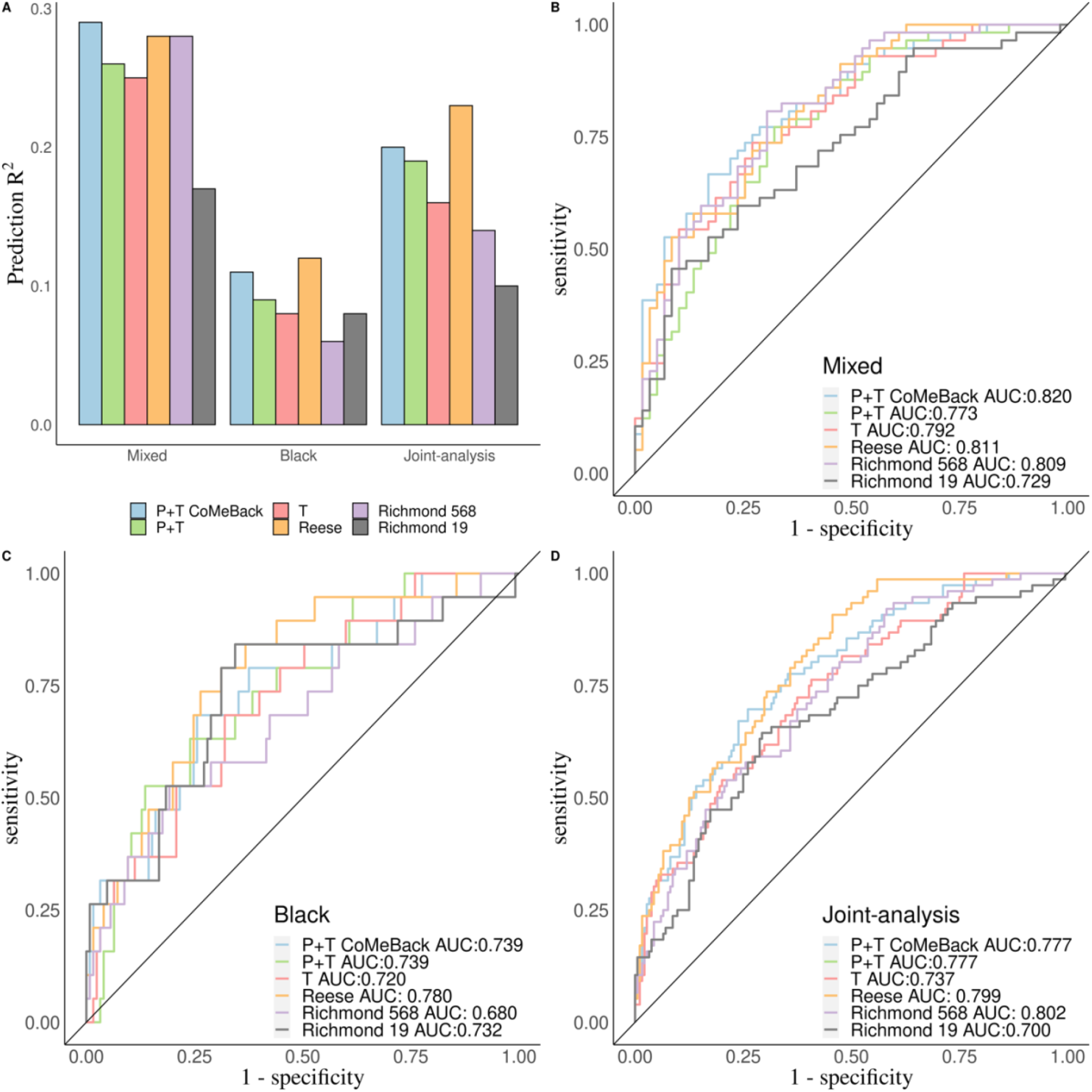
Real data application. Comparison of P+T CoMeBack method to P+T, T and 3 other published MRS for predicting maternal smoking status in the South African Drakenstein Child Health Study (DCHS). **A** Prediction R^2^ of all 6 MRS methods for Mixed infants, Black infants and pooled samples (joint-analysis). A receiver operating characteristic (ROC) curve comparing prediction performance of all 6 MRS among (**B**) Mixed infants, (**C**) Black infants and (**D**) pooled samples (joint-analysis).

**Table 2.** Association between maternal smoking status and MRS in DCHS.

| MRS | Mixed |  |  | Black |  |  | Pooled (Joint-analysis) |  |  |
| --- | --- | --- | --- | --- | --- | --- | --- | --- | --- |
|  | Beta-coefficient* | Standard Error | P-value | Beta-coefficient | Standard Error | P-value | Beta-coefficient | Standard Error | P-value |
| P+T CoMeBack MRS | 0.88 | 0.12 | 5.65 x10<sup>-11</sup> | 0.76 | 0.18 | 5.74 x10<sup>-5</sup> | 0.82 | 0.10 | 3.01x10<sup>-14</sup> |
| P+T MRS | 0.71 | 0.11 | 1.05 x10<sup>-8</sup> | 0.52 | 0.14 | 3.29 x10<sup>-4</sup> | 0.69 | 0.10 | 1.32x10<sup>-10</sup> |
| T MRS | 0.80 | 0.13 | 7.35 x10<sup>-9</sup> | 0.70 | 0.19 | 2.82 x10<sup>-4</sup> | 0.64 | 0.09 | 2.55 x10<sup>-11</sup> |
| Reese MRS | 0.97 | 0.13 | 3.77 x10<sup>-11</sup> | 0.73 | 0.16 | 2.06 x10<sup>-5</sup> | 0.77 | 0.09 | 2.50 x10<sup>-15</sup> |
| Richmond 568 MRS | 0.85 | 0.14 | 9.84 x10<sup>-9</sup> | 0.54 | 0.16 | 7.82 x10<sup>-4</sup> | 0.65 | 0.10 | 1.57 x10<sup>-10</sup> |
| Richmond 19 MRS | 0.76 | 0.14 | 5.37E x10<sup>-7</sup> | 0.70 | 0.19 | 3.78 x10<sup>-4</sup> | 0.70 | 0.11 | 2.05 x10<sup>-9</sup> |

**\*Beta-coefficients indicate change in interquartile range (IQR) of MRS between smokers and non-smokers.**

## Discussion

Based on the well-established P+T CoMeBack framework in PRS, we developed P+T CoMeBack MRS, which aggregates EWAS signals and could potentially be used as a biomarker in association studies where single CpGs do not achieve significance^4,40,41^. The proposed P+T CoMeBack MRS approach uses CoMeBack for co-methylation pruning and evaluates multiple P-value thresholds to maximize prediction performance. Such MRS could potentially serve as a powerful dimension reduction approach for mediation and multi-omics integration analyses^4,40-43^ as well as biomarkers of individual disease risk in a clinical setting^44-46^.

Overall, our simulation studies demonstrated good performance of P+T CoMeBack MRS for predicting phenotypes of interest with good statistical power and well-controlled type 1 error. We demonstrated that the prediction accuracy of MRS reflects the variance of phenotype that is explained by DNAm. By accounting for inter-correlation between CpGs, P+T CoMeBack MRS and P+T without CoMeBack showed a slightly better performance than the standard T method. In the real data application, we observed the best prediction of maternal smoking status when using P+T CoMeBack, which confirms the usefulness of accounting for co-methylation and demonstrates the ability of CoMeBack to control for false discover of CMR and usefulness in constructing MRS^22^. However, we note that P+T CoMeBack MRS could still have poor prediction performance if the external EWAS is underpowered or subject to bias.

In the prediction of maternal smoking status, P+T CoMeBack MRS showed comparable performance to Reese MRS, which was derived using the LASSO method^37^. When predictors are highly correlated, LASSO typically selects one of the correlated predictors and shrinks the effect size of the rest to zero, which might produce similar results to our pruning procedure in developing MRS. One of the advantages of P+T CoMeBack MRS is that it is based on EWAS summary statistics which are often publicly available, hence making it a valuable approach, as it is often difficult to obtain individual DNAm data from an external cohort. Additionally, P+T CoMeBack MRS can make use of meta-analysis-type summary statistics, which aggregates results from multiple studies to improve association estimates. In contrast, to construct MRS like Reese MRS, individual DNAm data are usually required to perform a LASSO regression, and these are often not accessible. Recently, novel penalized regressions have been proposed to generate PRS with only GWAS summary statistics and publicly available reference data^47^, but their applications to EWAS summary statistics for MRS have not been investigated. To develop MRS for different exposures and outcomes, we urge EWAS studies to make their genome-wide summary statistics publicly available.

In our simulation studies, weights obtained from Indian training samples were applied to generate P+T CoMeBack MRS, thus MRS among Indian testing samples were assumed to have the best prediction of the simulated phenotypes. However, MRS among Whites and Blacks also achieved a prediction accuracy as high as among Indians for both simulated phenotypes suggesting that genetic ancestry does not contribute to difference in prediction abilities of MRS across multi-ancestry population. This is likely because we assumed all ancestries share the same causal CpGs and effect sizes. However, in the real world, this assumption could possibly be violated for many phenotypes. Unlike ancestry in our simulation studies, ancestry in the real world is complex. The meaning of ancestry could be different in different regions/nations, and “effect of ancestry” involves the joint effects of ancestry-associated social determinants of health and environmental effects, and cultural context^48^. Ancestry, along with environment and social differences associated with it, could affect both MRS and phenotypes in numerous causal pathways and potentially modify the effect of MRS on the phenotypes. Thus, even if all ancestries indeed share the same causal CpGs and effect sizes, it might still not be sufficient to disentangle the relationship between ancestry, DNAm and phenotype of interest. This may greatly impact the transferability of MRS across different ancestries, which could be the reason why we observed an inconsistency of performance of P+T CoMeBack MRS in terms of their distributions and predictions across multi-ancestry population in the real data analyses. In practice, we recommend that researchers conduct MRS analyses stratified by ancestry first and evaluate the effect of ancestry on MRS analyses before pooling participants together for a joint analysis.

In our real data application, summary statistics for smoking were obtained from a cohort with mainly people of European ancestry^36^. MRS of smoking among mixed ancestry infants achieved a prediction accuracy of nearly 30%. However, the prediction accuracy of P+T CoMeBack MRS among Black African infants was only 10.9%. We suspect that the difference was largely due to the prevalence of active smoking among mothers of Black African infants being lower than those of mixed ancestry infants (13% vs 49%), which is similar to how the prevalence of outcome affects the predictive ability of PRS^49^. To the best of our knowledge, this is the first study to propose using CoMeBack for pruning MRS among multi-ancestry populations. However, there are several potential limitations that warrant mention. First, the sample size of both simulation studies and real data analyses was relatively small, thus our results might not fully capture the strengths and limitations of P+T CoMeBack MRS. Second, lack of different ancestry-specific summary statistics made it impossible to compare the use of external weights from population of different ancestries (e.g. European ancestry vs other ancestries). Third, the prevalence of active maternal smoking is different in different ancestries and has influenced the performance of P+T CoMeBack MRS. As a result, real data analysis of smoking MRS could not provide firm evidence about the transferability of MRS between Black African and mixed ancestry infants. Fourth, we mainly focused on the prediction performance of P+T CoMeBack MRS. Further studies are needed to assess the performance of P+T CoMeBack MRS in mediation analysis.

In conclusion, P+T in general and P+T using CoMeBack in particular, provides an improvement for prediction of phenotype of interest, over T method that does not account for co-methylation between CpGs. In contrast to PRS, using existing summary statistics that were derived from European populations can be used to calculate MRS in other ancestries, thus reducing the ancestry/ethnicity disparity in medical research. However, caution is needed in the analyses and interpretation of MRS results across multi-ancestry populations. More investigations of MRS are urged to further improve their prediction accuracy and translational values, also in combination with other clinical and non-clinical variables, especially among multi-ancestry population. With the current increase of large consortia-led EWAS for different exposures and health outcomes (e.g., the PACE consortium), we believe the predictive performance of MRS will continue to increase, and the P+T CoMeBack method has the potential to be widely used for risk prediction and association testing.

## Data Availability

All data produced in the present study are available upon reasonable request to the authors

## Acknowledgments

The authors thank the study and clinical staff at Paarl Hospital, Mbekweni and TC Newman clinics, as well as the CEO of Paarl Hospital, and the Western Cape Health Department for their support of the study. The authors thank the families and children who participated in this study.

## Notes

**Funding:** The Drakenstein Child Health Study was funded by the Bill & Melinda Gates Foundation (OPP 1017641), Discovery Foundation, South African Medical Research Council, National Research Foundation South Africa, CIDRI Clinical Fellowship and Wellcome Trust (204755/2/16/z). Additional support for the DNA methylation work was by the Eunice Kennedy Shriver National Institute of Child Health and Human Development of the National Institutes of Health (NICHD) under Award Number R21HD085849, and the Fogarty International Center (FIC). The content is solely the responsibility of the authors and does not necessarily represent the official views of the National Institutes of Health. AH is supported the HERCULES Center (NIEHS P30ES019776). DJS and HJZ are supported by the South African Medical Research Council (SAMRC). The funders had no role in the study design, data collection and analysis, decision to publish, or preparation of manuscript.

### Competing Interest Statement

The authors have declared no competing interest.

### Funding Statement

The Drakenstein Child Health Study was funded by the Bill & Melinda Gates Foundation (OPP 1017641), Discovery Foundation, South African Medical Research Council, National Research Foundation South Africa, CIDRI Clinical Fellowship and Wellcome Trust (204755/2/16/z). Additional support for the DNA methylation work was by the Eunice Kennedy Shriver National Institute of Child Health and Human Development of the National Institutes of Health (NICHD) under Award Number R21HD085849, and the Fogarty International Center (FIC). The content is solely the responsibility of the authors and does not necessarily represent the official views of the National Institutes of Health. AH is supported the HERCULES Center (NIEHS P30ES019776). DJS and HJZ are supported by the South African Medical Research Council (SAMRC). The funders had no role in the study design, data collection and analysis, decision to publish, or preparation of manuscript.

### Author Declarations

The DCHS staff obtain written consent from mothers on an annual basis and the study was approved by the Ethics Committee of the Faculty of Health Sciences, University of Cape Town, by Stellenbosch University and the Western Cape Provincial Research committee

### Summary of Updates

We have included P+T method without CoMeBack in our simulation studies and real data application for comparison.

