## SupplementalFigures for "Pruning and thresholding approach for methylation risk scores in multi-ancestry populations"

### Supplementary Figures

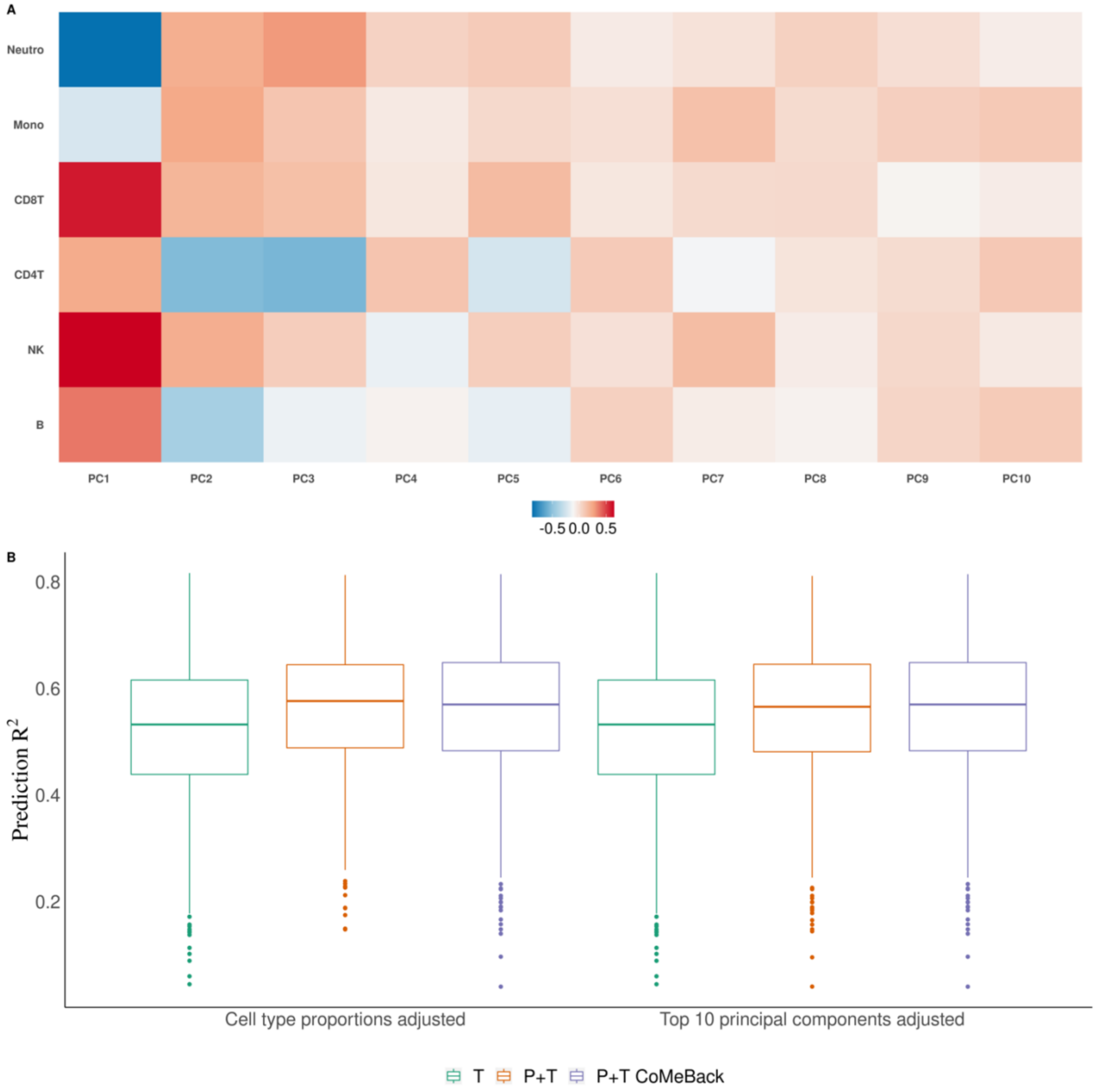


**Supplement figure 1. Comparison between adjustment for cell type proportions and adjustment for top 10 principal components in simulation studies. (A)** Heatmap showing correlation between cell type proportions and top 10 principal components in simulation dataset. (B) Prediction $R^2$ of P+T CoMeBack, P+T and T method using summary statistics adjusted for cell type proportions or top 10 principal components, among Indian participants.

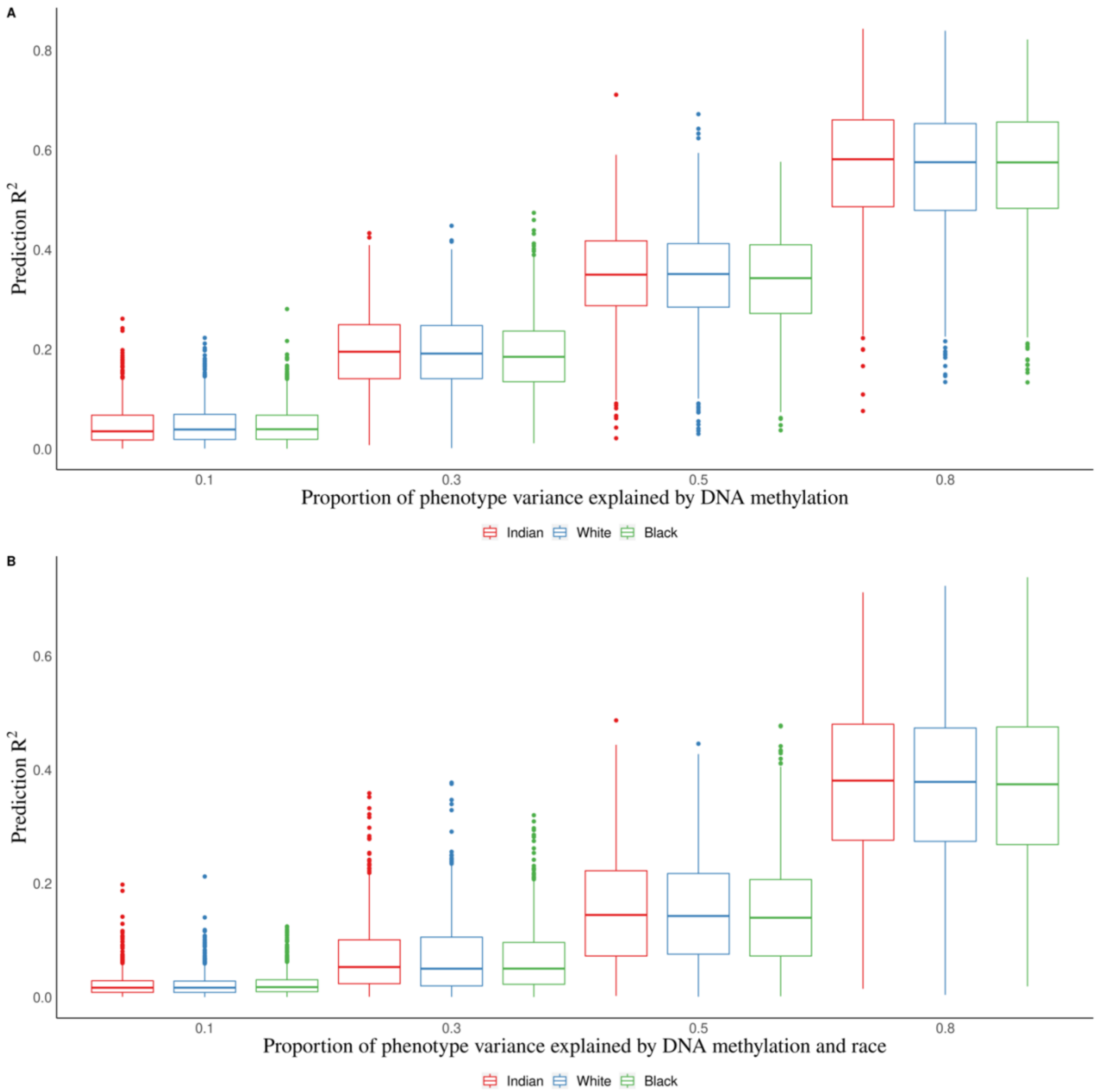


**Supplement figure 2. Prediction $R^2$ of P+T CoMeBack method among multi-ancestry populations in simulation scenarios with different proportions of phenotype variance explained by DNA methylation.** For each simulation, the discovery cohort was repeatedly and randomly split into a training set comprising 762 Indians and a testing set comprising 136 people of each ancestry group. The proportion of causal CpGs located in CMR is 70%. Results are shown for the prediction of simulated phenotypes (**A**) without an influence of ancestry and (**B**) influenced by ancestry. Each box represents the distribution of prediction accuracy across 1000 simulations, where the central mark is the median, the edges of the box are the $25^{th}$ and $75^{th}$ percentiles.

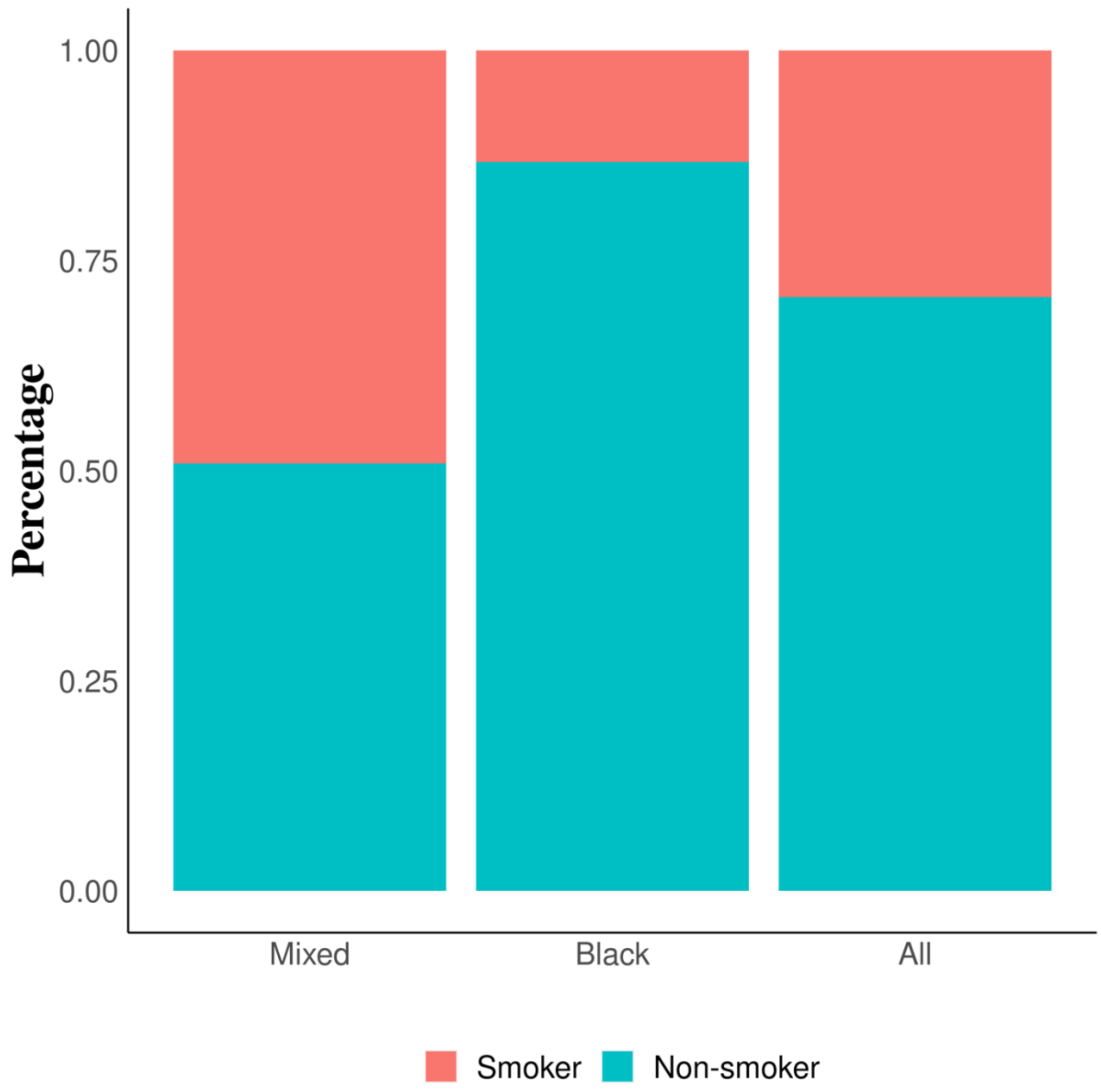


**Supplement figure 3. Prevalence of non-smokers, passive smokers & active smokers among Mixed infants, Black infants and pooled samples in the South African Drakenstein Child Health Study (DCHS).**

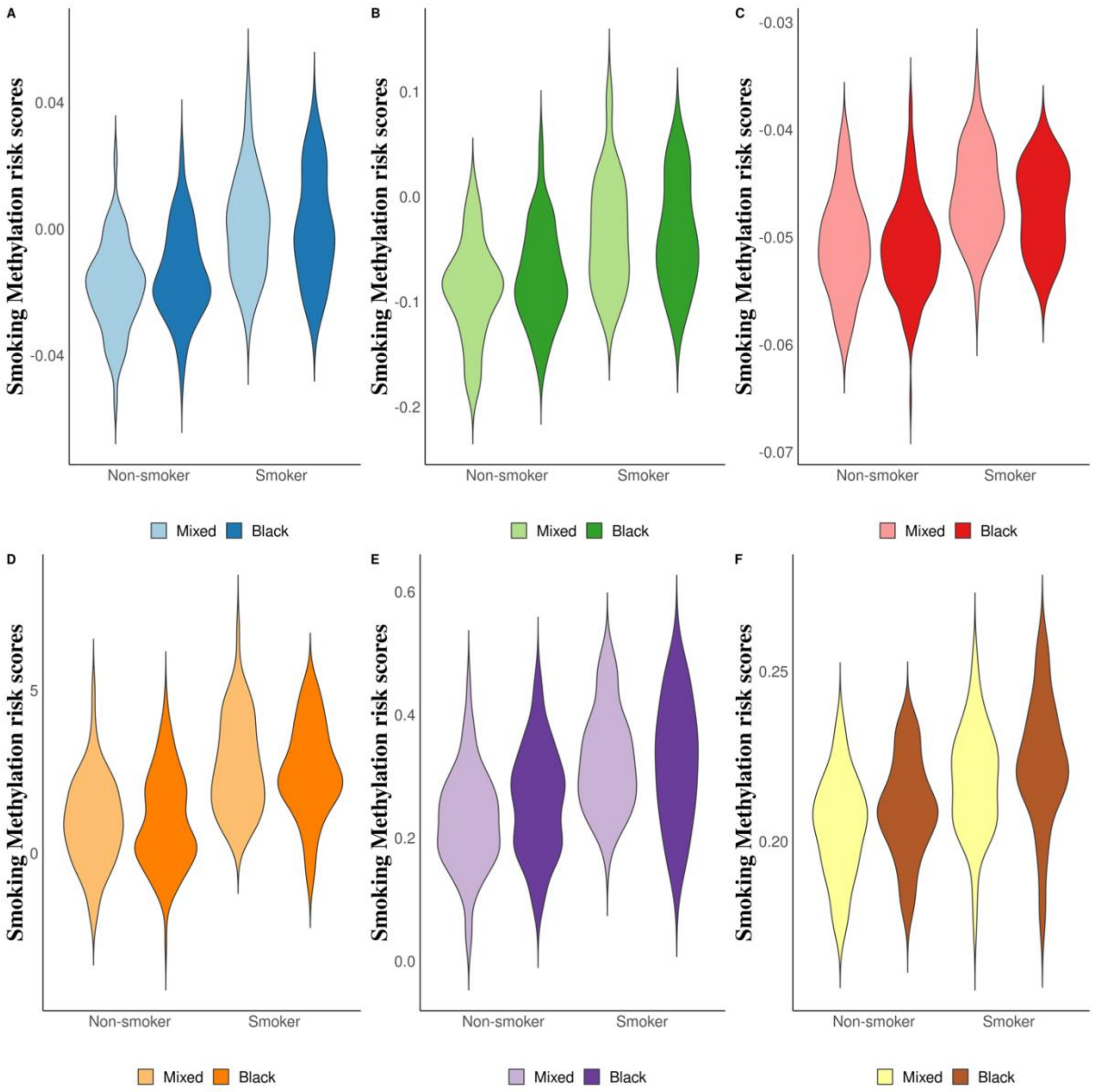


**Supplement figure 4. Real data application. Comparison of P+T CoMeBack method to P+T, T and 3 other published MRS for predicting maternal smoking status in the South African Drakenstein Child Health Study (DCHS).** Distribution of (**A)** P+T CoMeBack MRS (**B**) P+T MRS, (**C)** T MRS, (**D**) Reese MRS, (**E)** Richmond 568 MRS and (**F**) Richmond 19 MRS among non-smokers and smokers stratified by ancestry.
